# Comparison of Relapse Rate and Disease Severity among patients with Type 2 Lepra Reaction receiving Tofacitinib and Thalidomide separately as an adjuvant to systemic steroids: A Longitudinal Analytical Study

**DOI:** 10.64898/2026.07.07.26357443

**Authors:** Rishav Sanghai, Bijaya Nanda Naik, Rakhee Gupta, Gaurav Dash, Irene Mathews, Swetalina Pradhan

## Abstract

**Background:** Erythema nodosum leprosum (ENL) is a severe immune-mediated complication of multibacillary leprosy requiring prolonged immunosuppression. Steroid-sparing agents are essential to reduce relapse and treatment-related morbidity.

**Methods:** This longitudinal analytical observational study compared outcomes in patients with ENL treated with prednisolone plus thalidomide (Group A; n=30) and prednisolone plus tofacitinib (Group B; n=31). Patients were followed for 6 months. Primary outcomes included relapse rate and ENLIST ENL Severity Score (EESS). Secondary outcomes were neutrophil–lymphocyte ratio (NLR), Dermatology Life Quality Index (DLQI), steroid dependency, and adverse events. Inter-group comparisons and longitudinal analyses were performed using non-parametric tests. Correlations between NLR, EESS, and DLQI were assessed using Spearman’s rank correlation.

**Results:** Relapse occurred in 36.7% of patients in Group A and 71.0% in Group B (p=0.007). The mean number of relapses was significantly lower in Group A (0.70±1.06 vs 1.84±1.51; p=0.002). At 3 and 6 months, Group A demonstrated significantly lower NLR values (p=0.017 and p<0.001, respectively). DLQI and EESS scores improved in both groups; however, sustained improvement was more consistent in Group A. Steroid-free status at 6 months was achieved in 93.3% of Group A compared with 58.1% of Group B (p<0.001). NLR showed a positive correlation with EESS (ρ=0.269, p=0.018) and DLQI (ρ=0.604, p<0.001) at 6 months. On multivariable logistic regression analysis adjusting for baseline confounders, patients receiving tofacitinib had significantly higher odds of relapse compared with those receiving thalidomide (adjusted OR 9.87, 95% CI 1.73–27.12; p = 0.006).Adverse events were predominantly mild to moderate, with differing safety profiles between groups.

**Conclusion:** Thalidomide demonstrated superior relapse prevention and steroid-sparing efficacy compared with tofacitinib in ENL. NLR correlated with disease severity and quality of life, supporting its role as a useful biomarker for monitoring disease activity during follow-up.

## Introduction

Erythema nodosum leprosum (ENL), also referred to as Type 2 lepra reaction, is a complex immune-mediated inflammatory complication predominantly affecting patients with multibacillary leprosy. It is characterized by recurrent painful erythematous nodules accompanied by systemic manifestations such as fever, arthritis, neuritis, lymphadenitis, and visceral organ involvement, leading to significant morbidity and impaired quality of life. In India, where multibacillary leprosy remains prevalent, ENL continues to pose a major therapeutic challenge.^1,2^

Systemic corticosteroids constitute the mainstay of treatment for moderate to severe ENL because of their rapid anti-inflammatory effects. However, prolonged or recurrent corticosteroid use is associated with substantial adverse effects, including metabolic derangements, infections, osteoporosis, and adrenal suppression, necessitating the search for effective steroid-sparing therapies.^3^

Thalidomide has long been regarded as one of the most effective adjuvant agents for ENL owing to its potent anti–tumour necrosis factor-α (TNF-α) activity and immunomodulatory effects.^4^ Despite its efficacy, its use is limited by serious adverse effects such as teratogenicity, peripheral neuropathy, and thromboembolic risk, prompting interest in alternative targeted immunomodulatory therapies.

Tofacitinib, an oral Janus kinase (JAK-1 and JAK-3) inhibitor, modulates multiple cytokine signalling pathways implicated in ENL pathogenesis, including TNF-α, interleukin-6, interleukin-17, and interferon-γ.^5^ In recent years, case reports and small case series have described its successful use in refractory ENL and severe atypical lepra reactions, including Lucio phenomenon.^6–8^ However, evidence supporting its use is limited to isolated reports, and comparative data evaluating its efficacy against established therapies such as thalidomide remain scarce.

There is also growing interest in identifying simple and accessible biomarkers that reflect disease activity and systemic inflammation in ENL. The neutrophil–lymphocyte ratio (NLR), derived from routine complete blood counts, has been proposed as a useful marker of disease severity and prognosis in ENL.^9–11^

Against this background, the present study was undertaken to compare relapse rates, disease severity, systemic inflammatory burden, quality of life, and safety outcomes between tofacitinib and thalidomide when used as adjuvants to systemic corticosteroids in patients with ENL.

## MATERIALS AND METHODS

### Study Design and Setting

The study was commenced after approval from the Institute Research Committee and Institute Ethics Committee. This was a longitudinal analytical observational study with a historical comparison group, conducted at a tertiary care referral centre for Hansen disease in eastern India. The study was designed to compare clinical outcomes in patients with Type 2 lepra reaction (erythema nodosum leprosum, ENL) treated with prednisolone in combination with either thalidomide or tofacitinib over a follow-up period of six months.

### Study Period

Patients treated with prednisolone and thalidomide were identified retrospectively from January 2022 to December 2023. Patients treated with prednisolone and tofacitinib were prospectively enrolled between January 2024 and December 2025. The study commenced on October 2024 and last patient was recruited till November 2025, to have completed follow-up latest by May 2026.

### Inclusion and Exclusion Criteria

Adult patients aged 18 years or older with a clinical diagnosis of Type 2 lepra reaction belonging to the multibacillary spectrum of Hansen disease (borderline lepromatous or lepromatous leprosy) were eligible for inclusion. Only patients who completed follow-up visits at 1 month, 3 months, and 6 months were included in the analysis. Patients were excluded if they were pregnant or lactating, had active systemic infections including tuberculosis, severe hepatic or renal dysfunction, significant haematological abnormalities, active neuritis requiring alternative immunosuppressive therapy, known hypersensitivity to study medications, or incomplete follow-up.

### Study Population

During the study period, clinical records of 1,073 patients diagnosed with Hansen disease were screened. Among these, 164 patients were identified as having Type 2 lepra reaction. After applying predefined inclusion and exclusion criteria and excluding patients with incomplete follow-up, a total of 61 patients who completed all scheduled follow-up visits were included in the final analysis. These patients were divided into two treatment groups: Group A consisted of 30 patients who received prednisolone with thalidomide, and Group B consisted of 31 patients who received prednisolone with tofacitinib as shown **in Figure 1**.

**Figure 1:**
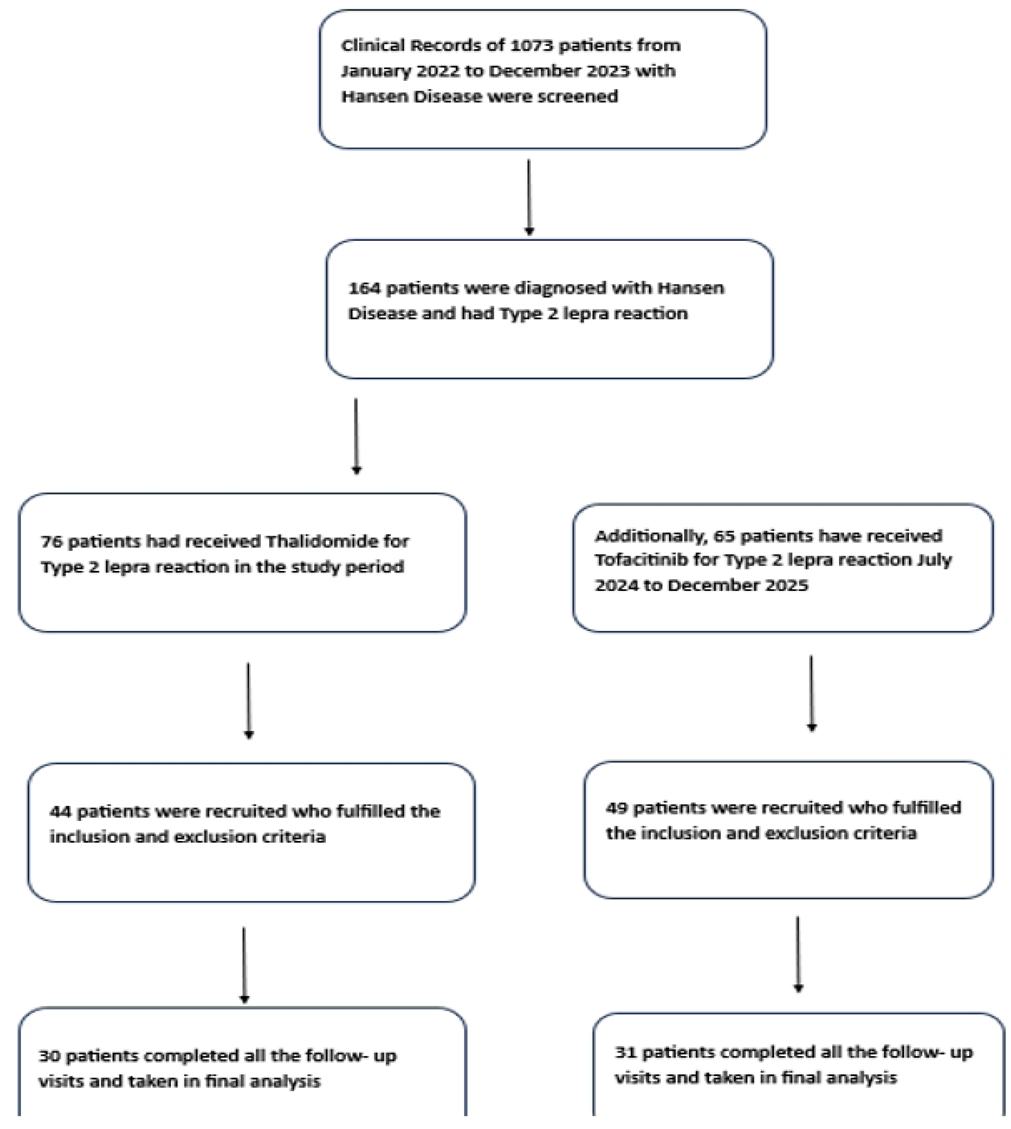
Participant Flow of the Study – Demonstrating flow of patient in Thalidomide and Tofacitinib Arm.

### Sample Size Calculation

Sample size estimation was based on relapse rate as the primary outcome. Previous studies evaluating thalidomide in erythema nodosum leprosum have reported recurrence rates of approximately 15–20%. As comparative data for tofacitinib were unavailable, a clinically meaningful difference of 35% between treatment groups was assumed. Using a two-sided alpha of 0.05 and power of 80%, the minimum estimated sample size was 28 patients per group.

### Study Population and Participant Flow

During the study period from **January 2022 to December 2023**, clinical records of **1,073 patients with Hansen disease** were screened. Among these, **164 patients** were diagnosed with Type 2 lepra reaction (erythema nodosum leprosum, ENL). After applying predefined inclusion and exclusion criteria and excluding patients with incomplete follow-up, **61 patients** who completed all scheduled visits were included in the final analysis for Thalidomide group. Similarly, 65 patients received Prednisolone and Tofacitinib for Type 2 Lepra reaction from January 2024 till December 2025. Out of them 49 patients fulfilled the inclusion and exclusion criteria. Finally, 31 patients were recruited for final analysis after having completed all follow up visits.

These patients were divided into two treatment groups: **Group A (prednisolone + thalidomide; n = 30)** and **Group B (prednisolone + tofacitinib; n = 31)**. The study enrollment and attrition process is depicted in **Figure 1**.

### Baseline Assessment

At enrolment, detailed demographic and socioeconomic data including age, sex, residence, and socioeconomic status based on the modified Kuppuswamy scale were recorded. Disease-related variables such as clinical spectrum according to the Ridley–Jopling classification, duration of leprosy, treatment status, and slit skin smear results with bacteriological index were documented. The clinical pattern of ENL was categorized as recurrent or chronic. Baseline disease severity was assessed using the ENLIST ENL Severity Score (EESS). Systemic inflammatory status was assessed using the neutrophil–lymphocyte ratio (NLR), calculated from complete blood count parameters. Dermatology-specific quality of life was assessed using the Dermatology Life Quality Index (DLQI).

### Treatment Protocol

All patients received systemic corticosteroid therapy in the form of prednisolone according to institutional treatment guidelines. In Group A, thalidomide was administered as an adjuvant agent along with prednisolone, whereas in Group B, tofacitinib was used as the adjuvant therapy. Dose adjustments and tapering of corticosteroids were guided by clinical response and treating physician discretion.

### Follow-up and Outcome Measures

Patients were followed up at 1 month, 3 months, and 6 months. At each visit, disease severity was reassessed using EESS, systemic inflammatory status was evaluated using NLR, and quality of life was assessed using DLQI. Relapse events, defined as recurrence of ENL symptoms requiring escalation or reintroduction of systemic therapy, were recorded. Steroid dependency was assessed at the end of 6 months, defined as the inability to completely taper systemic corticosteroids. Adverse effects were actively monitored and documented at each follow-up visit.

### Statistical Analysis

Statistical analysis was performed using Microsoft Excel 2010 and Jamovi 2.0. Continuous variables were expressed as mean with standard deviation or median with interquartile range depending on data distribution, while categorical variables were expressed as frequencies and percentages. Inter-group comparisons were conducted using the Mann–Whitney U test for continuous variables and the chi-square test for categorical variables. Longitudinal intra-group comparisons across follow-up visits were analyzed using the Friedman test followed by post-hoc Durbin–Conover pairwise comparisons where applicable. Correlations between neutrophil–lymphocyte ratio, ENLIST ENL Severity Score, and Dermatology Life Quality Index were assessed using Spearman’s rank correlation coefficient. A p-value of less than 0.05 was considered statistically significant.

### Ethics Statement

This study was conducted in accordance with the ethical principles of the Declaration of Helsinki. Ethical approval was obtained from the Institutional Ethics Committee of AIIMS Patna before commencement of the study (Approval No.: AIIMS/Pat/IEC/2024/PGTh/Jan24/PG47; Date: _5/11/24). Written informed consent was obtained from all participants prior to their enrollment in the study. Participants were informed about the objectives, procedures, potential benefits, and possible risks of the study, and confidentiality of patient information was maintained throughout the study.

### Data Availability Statement

The datasets generated and analyzed during the current study contain sensitive patient information and are not publicly available due to institutional ethics and patient confidentiality restrictions. De-identified data may be made available from the corresponding author upon reasonable request and with permission from the Institutional Ethics Committee.

## RESULTS

A total of 61 study subjects were recruited after applying the pre-defined inclusion and exclusion criteria. Out of them 30 patients were in Group A received Thalidomide & Prednisolone and 31 patients were in Group B, received Tofacitinib & Prednisolone.

The overall mean age of the study population was 35.0 ± 10.8 years (range: 18–60 years), with comparable age distribution between Group A (35.8 ± 10.4 years) and Group B (34.1 ± 11.4 years). Male patients constituted 80.3% (49/61) of the cohort, with similar sex distribution across both groups. Most participants resided in rural areas (77.0%), followed by semiurban (16.4%) and urban (6.6%) settings. According to the modified Kuppuswamy socioeconomic scale, the majority belonged to the upper-lower socioeconomic class (60.7%), with no statistically significant differences between treatment groups.

Baseline demographic and socioeconomic characteristics were well balanced between Group A and Group B, as detailed in Table 1.

**Table 1:** Baseline demographic and disease-related characteristics of patients with ENL in Group A and Group B.

**Figure 9g . One of our study subject having resolved necrotic ENL the forearm – resolving with atrophic scar**
|  |  | Group A (n=30) | Group B (n=31) |
| --- | --- | --- | --- |
| Age and Sex | Mean Age in years (SD) | 35.8 (10.4) | 34.1(11.4) |
|  | Male | 25(83.3%) | 24(77.4%) |
|  | Female | 5(16.7%) | 7(22.6%) |
| Socioeconomic Status | Lower Class | 3(10%) | 4(12.9%) |
|  | Upper Lower Class | 15(50%) | 22(71%) |
|  | Modified Kuppuswamy Score (SD) | 10.2(4.7) | 8.1(3.7) |
| Disease Profile | LL Hansen Disease | 28 (93.3%) | 26(83.9%) |
|  | BL Hansen Disease | 2(6.7%) | 5(16.1%) |
|  | Duration of Disease (SD) in months | 20.6(14.5) | 25.5(15.5) |
|  | Treatment Naïve | 11(36.7%) | 11(35.5%) |
|  | Released From Treatment | 5(16.7%) | 3(9.8%) |
| Slit Skin Smear | 4+ and Above | 25(83.3%) | 26(83.9%) |
|  | 3+ and below | 5(16.7%) | 5(16.1%) |
| Outcome Variables | EES Score (SD) | 16.4(4.7) | 17.4(3.5) |
|  | Neutrophil Lymphocyte Ratio | 6.6(3.5) | 6.3(3.5) |
|  | DLQI | 15.2(4.8) | 16.2(4.0) |

The patients were divided into clinical spectrum of Hansen Disease as per Ridely Jopley classification and WHO criteria. It was seen that 88.4% patients were in Lepromatous Leprosy (LL) and 11.6% patients were in Borderline Lepromatous (BL) spectrum. All patients in our study were in the Multibacillary pole of Hansen Disease (100%).

The mean duration of leprosy disease before presentation was 20.57 ± 14.46 months among patients treated with prednisolone and thalidomide, whereas patients receiving prednisolone and tofacitinib had a longer mean disease duration of 25.52 ± 15.50 months.

In Slit skin Smear, the distribution of bacteriological index (BI) grades showed a marked predominance of high BI positivity. Overall, 51 out of 61 patients (83.6%) had a high bacillary load (BI ≥4+), indicating that the majority of the study population had multibacillary disease. BI 5+ was the most frequent finding, observed in 24 patients (39.3%), followed by BI 4+ in 16 patients (26.2%) and BI 6+ in 11 patients (18.0%). No patients had negative slit skin smear results (0%). The distribution of BI grades was comparable between Group A and Group B, with both groups showing clustering in the higher BI categories. The distribution of patients according to treatment status shows that the largest proportion were treatment-naïve, comprising 22 patients (approximately 36%) of the total study population. This was followed by 20 patients (about 33%) who were on multidrug therapy (MDT) for less than one year, indicating that a substantial proportion of cases were either newly diagnosed or in the early phase of treatment. 11 patients (around 18%), had completed more than one year but less than two years of MDT, reflecting progression through the treatment course. The smallest group consisted of 8 patients (approximately 13%) who had been released from treatment, suggesting a comparatively lower number of patients who had completed therapy at the time of assessment.

Recurrent ENL was the most common clinical pattern, observed in 62.3% of patients, while 37.7% had chronic ENL.

At baseline, disease severity assessed by the ENLIST ENL Severity Score (EESS) was similar between Group A (16.4 ± 4.7) and Group B (17.4 ± 3.5). Baseline neutrophil-to-lymphocyte ratio (NLR) was also comparable (Group A: 6.65 ± 3.54; Group B: 6.29 ± 3.59), indicating similar systemic inflammatory status.Baseline Dermatology Life Quality Index (DLQI) scores reflected severe quality-of-life impairment in both groups (Group A: 15.2 ± 4.8; Group B: 16.2 ± 4.0). These findings confirm that both groups were clinically and biologically comparable at enrolment. (Table 1).

Relapse during the 6-month follow-up period occurred in 11 of 30 patients (36.7%) in Group A and 22 of 31 patients (71.0%) in Group B. This difference was statistically significant (χ² = 7.22, df = 1, p = 0.007), demonstrating a significantly higher relapse risk in patients receiving prednisolone plus tofacitinib. In addition to relapse frequency, relapse burden differed significantly between groups. The mean number of relapse episodes was 0.70 ± 1.06 in Group A compared with 1.84 ± 1.51 in Group B. Median relapse counts were 0 and 2, respectively. This difference was statistically significant on Mann–Whitney U testing (U = 263, p = 0.002). Relapse outcomes are summarized in Table 2 and illustrated in **Figure 2**. At the end of 6 months, 28 of 30 patients (93.3%) in Group A were successfully weaned off systemic corticosteroids, compared with 18 of 31 patFigurients (58.1%) in Group B. Conversely, 41.9% of patients in Group B remained steroid-dependent versus 6.7% in Group A. The odds of remaining steroid-dependent were significantly lower in Group A (OR = 0.099; 95% CI: 0.020–0.491; p < 0.001). Steroid dependency outcomes are presented in **Figure 3**.

**Figure 2.**
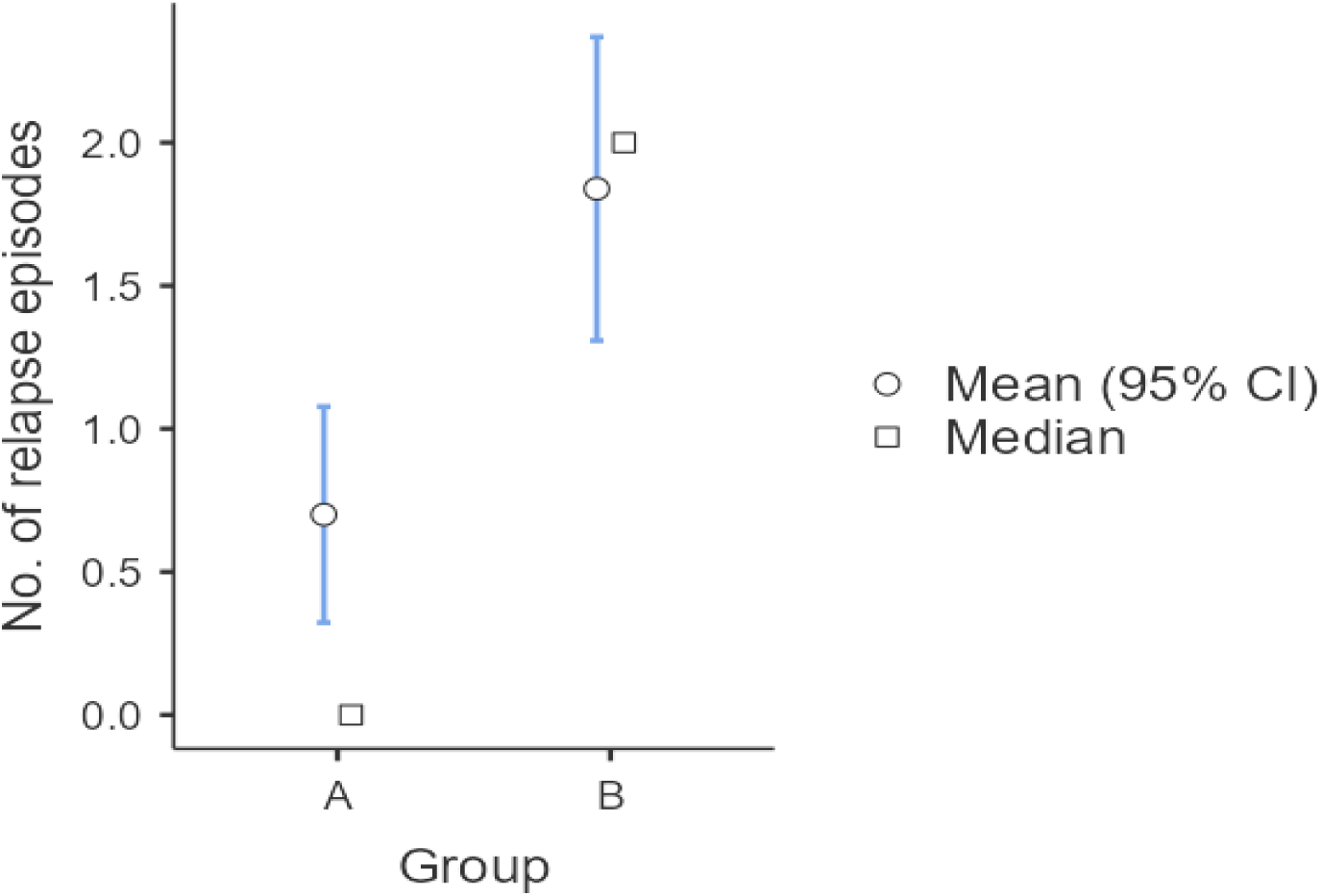
Comparison of mean number of relapse episodes between Group A and Group B during the follow-up period.

**Figure 3.**
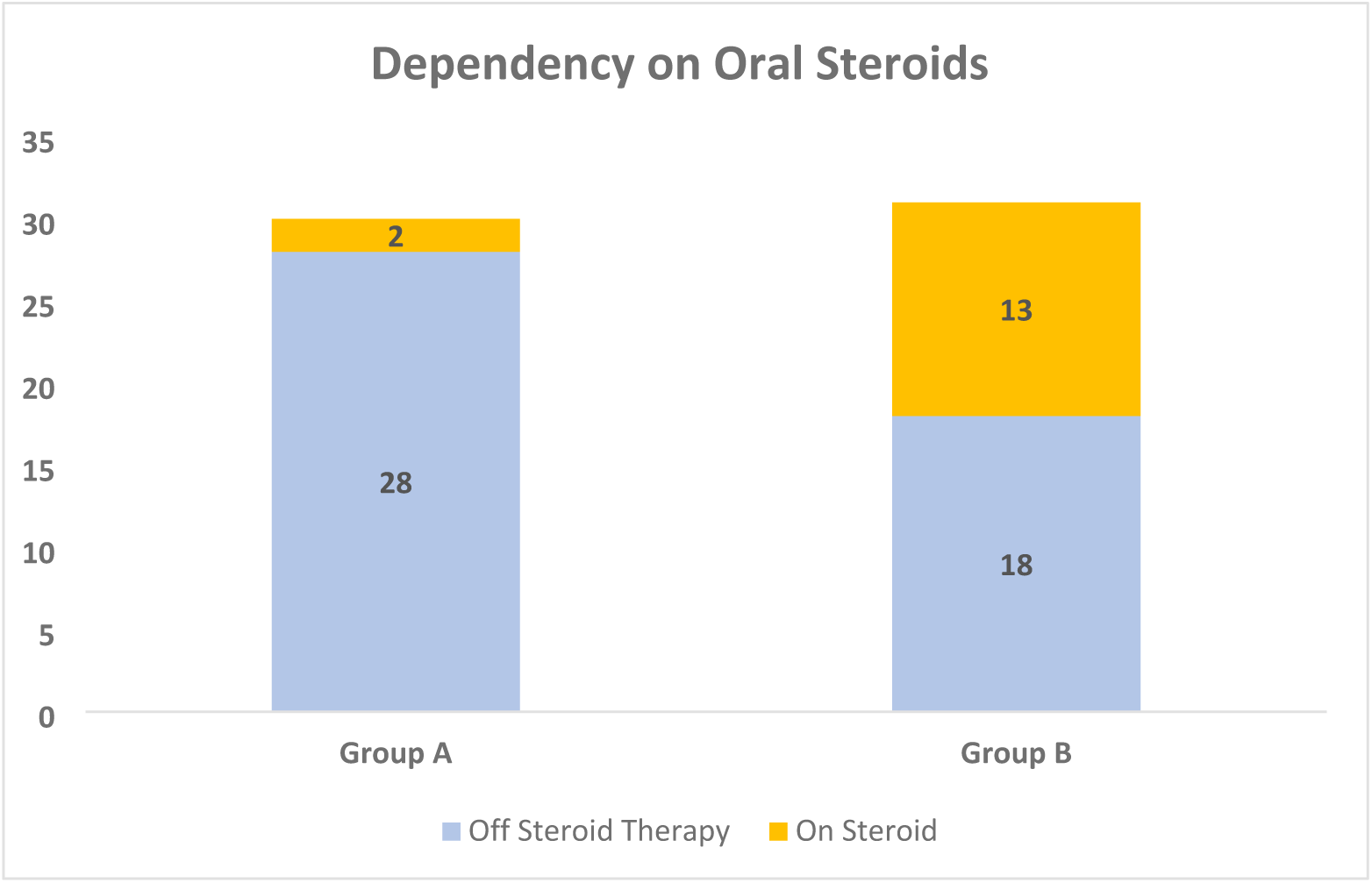
Comparison of steroid dependency between Group A and Group B during the follow-up period.

**Table 2:** Comparison of Outcome variables in Group A and Group B (with level of significance)

|  |  | Group A (n=30) | Group B (n=31) | P- value |
| --- | --- | --- | --- | --- |
| ENLIST ENL Severity Score | 1 month | 6.0 | 6.0 | 0.948 |
|  | 3 month | 3.0 | 9.0 | 0.002 |
|  | 6 months | 1.73 | 2.0 | 0.072 |
| Neutrophil-Lymphocyte Ratio | 1 month | 5.15 | 4.50 | 0.516 |
|  | 3 month | 4 | 4.40 | 0.017 |
|  | 6 months | 2.75 | 4.50 | <0.001 |
| Dermatology Life Quality Index (DLQI) | 1 month | 8.0 | 7.0 | 0.229 |
|  | 3 months | 5 | 7.0 | 0.024 |
|  | 6 months | 3.0 | 9.0 | <0.001 |
| Relapse | Relapse Rate | 11(36.7%) | 22(71%) | 0.007 |
|  | Mean Number of Relapses (SD) | 0.7 (1.06) | 1.84(1.51) | 0.002 |
| Dependency on Steroids | Number of patients who were steroid free | 28(93.3%) | 18(58.1%) | <0.001 |
| Adverse Effects Profile (Instances including all follow-up visits) | Constipation | 27(30%) | 11(11.8%) | <0.002 |
|  | Diarrhoea, Nausea and Abdominal Pain | 4(4.5%) | 24(26%) | <0.001 |
|  | Transaminitis, Hyperlipidaemia | 3(3.6%) | 26(28%) | <0.001 |
|  | Headache, Drowsiness, Somnolence | 15(16.7%) | 2(2.5%) | 0.003 |
|  | Others | Congestion of Throat -1; Nasal Discharge -1 | Herpes Zoster -2 |  |

The secondary outcome measures of change in EESS score, NLR and DLQI were assessed in both the groups across the 6 month follow-up period. Mixed repeated-measures ANOVA demonstrated a significant effect of time on EESS scores (F(3,177)=172.72, p<0.001, η²=0.619), indicating that EESS scores changed significantly across the follow-up period. The large effect size suggests that approximately 61.9% of the variance in EESS scores was attributable to time.

A significant Time × Group interaction was also observed (F(3,177)=5.51, p=0.001, η²=0.020), indicating that the pattern of change in EESS scores over time differed significantly between the Prednisolone + Thalidomide group and the Prednisolone+Tofacitinib group. This suggests that the response to treatment over the study period was not uniform across the two treatment groups.

Post Hoc Analysis and Comparison of EESS scores between the two treatment groups revealed no significant difference at baseline (16.40 ± SE 0.76 vs 17.39 ± SE 0.74; p=0.982) or at 1 month (7.10 ± SE 0.99 vs 6.97 ± SE 0.98; p=1.000). However, at 3 months, Group 1 demonstrated a significantly lower EESS score compared with Group 2 (3.83 ± SE 0.88 vs 8.65 ± SE 0.87; p=0.006), indicating superior clinical improvement. By 6 months, EESS scores had decreased substantially in both groups and the difference between groups was no longer statistically significant (1.73 ± SE 0.11 vs 2.00 ± SE 0.11; p=0.681). It is also represented in Figure 4

**Figure 4.**
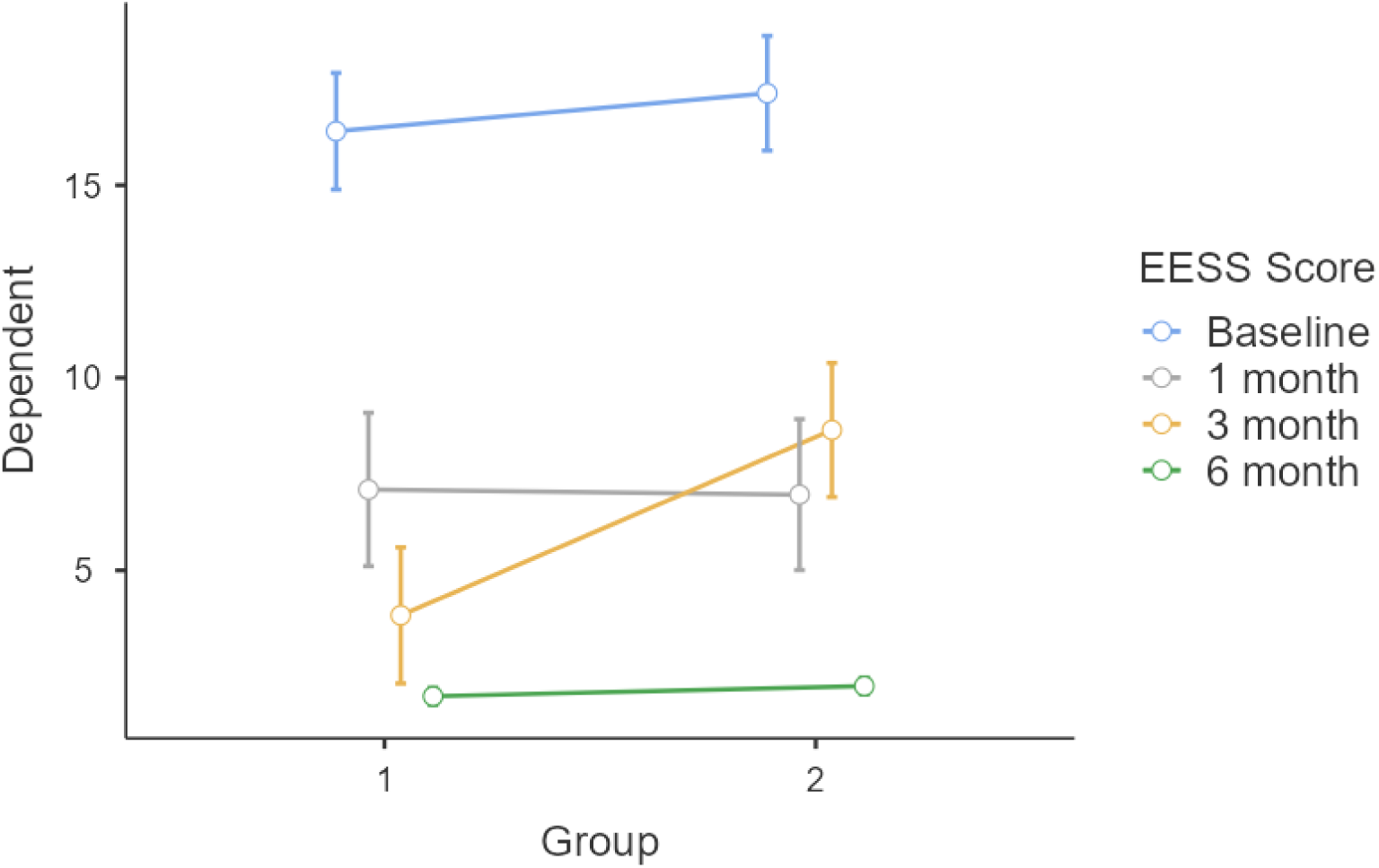
Comparison of EESS between Group A and Group B during the follow-up period at 1 month, 3 months and 6 months.

Similarly, Mixed repeated-measures ANOVA demonstrated a significant effect of time on NLR (F(3,177)=16.15, p<0.001, η²=0.142), indicating that NLR changed significantly during the follow-up period. The effect size suggests that approximately 14.2% of the variance in NLR was attributable to changes over time. A significant Time × Group interaction was also observed (F(3,177)=3.75, p=0.012, η²=0.033), indicating that the pattern of change in NLR over time differed significantly between the Prednisolone + Thalidomide group and the Prednisolone +Tofacitinib group. This finding suggests that the treatments exerted different effects on NLR during follow-up.

Post hoc comparison and analysis of means were done. The mean NLR decreased progressively in both treatment groups throughout the follow-up period. At baseline, there was no significant difference in NLR between the Prednisolone + Thalidomide group and the Prednisolone group (6.65 ± 0.65 vs 6.29 ± 0.64; p=1.000). Similarly, no significant difference was observed at 1 month (5.59 ± 0.41 vs 5.17 ± 0.40; p=1.000).

At 3 months, the Prednisolone + Thalidomide group demonstrated a lower mean NLR than the Prednisolone group (3.84 ± 0.40 vs 5.22 ± 0.39); however, the difference was not statistically significant after Bonferroni correction (p=0.443).

At 6 months, the Prednisolone + Thalidomide group showed a significantly lower NLR compared with the Prednisolone group (2.91 ± 0.25 vs 4.60 ± 0.24; p<0.001), indicating a greater reduction in systemic inflammation in the combination therapy arm as shown in Figure 5. Mixed repeated-measures ANOVA demonstrated a significant effect of time on NLR (F(3,177)=16.15, p<0.001, η²=0.142), indicating significant changes in NLR during follow-up. A significant Time × Group interaction was also observed (F(3,177)=3.75, p=0.012, η²=0.033), suggesting that the pattern of NLR change over time differed significantly between the two treatment groups.

**Figure 5.**
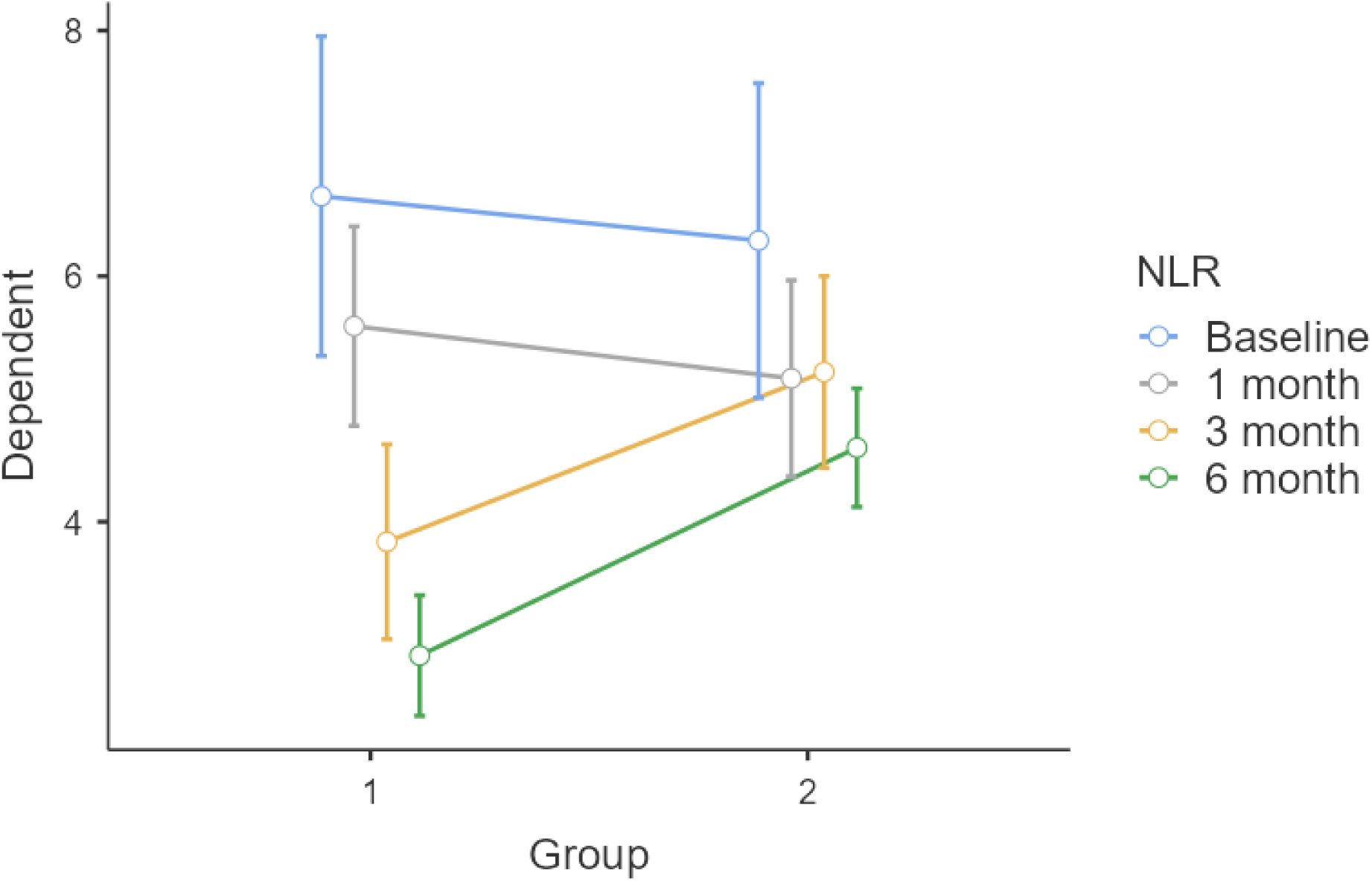
Comparison of Neutrophil-Lymphocyte Ratio between Group A and Group B during the follow-up period at 1 month, 3 months and 6 months.

Mixed repeated-measures ANOVA demonstrated a significant effect of time on DLQI scores (F(3,177)=76.14, p<0.001, η²=0.418), indicating that DLQI scores changed significantly during the follow-up period. The effect size suggests that approximately 41.8% of the variance in DLQI scores was attributable to changes over time.

A significant Time × Group interaction was also observed (F(3,177)=5.55, p=0.001, η²=0.031), indicating that the pattern of change in DLQI scores over time differed significantly between the Prednisolone + Thalidomide group and the Prednisolone group. This finding suggests that the impact of treatment on quality of life varied between the two treatment arms during follow-up.

DLQI scores decreased progressively in both treatment groups during follow-up, indicating improvement in quality of life. At baseline, DLQI scores were comparable between the Prednisolone + Thalidomide and Prednisolone groups (15.20 ± 0.81 vs 16.16 ± 0.80; p=0.989). Similarly, no significant differences were observed at 1 month (8.63 ± 0.79 vs 8.03 ± 0.77; p=0.999) or 3 months (5.67 ± 0.77 vs 8.29 ± 0.76; p=0.247).

At 6 months, the Prednisolone + Thalidomide group demonstrated significantly lower DLQI scores compared with the Prednisolone group (3.50 ± 0.80 vs 8.45 ± 0.78; p=0.001), indicating superior improvement in dermatology-specific quality of life as shown in Figure 6

**Figure 6.**
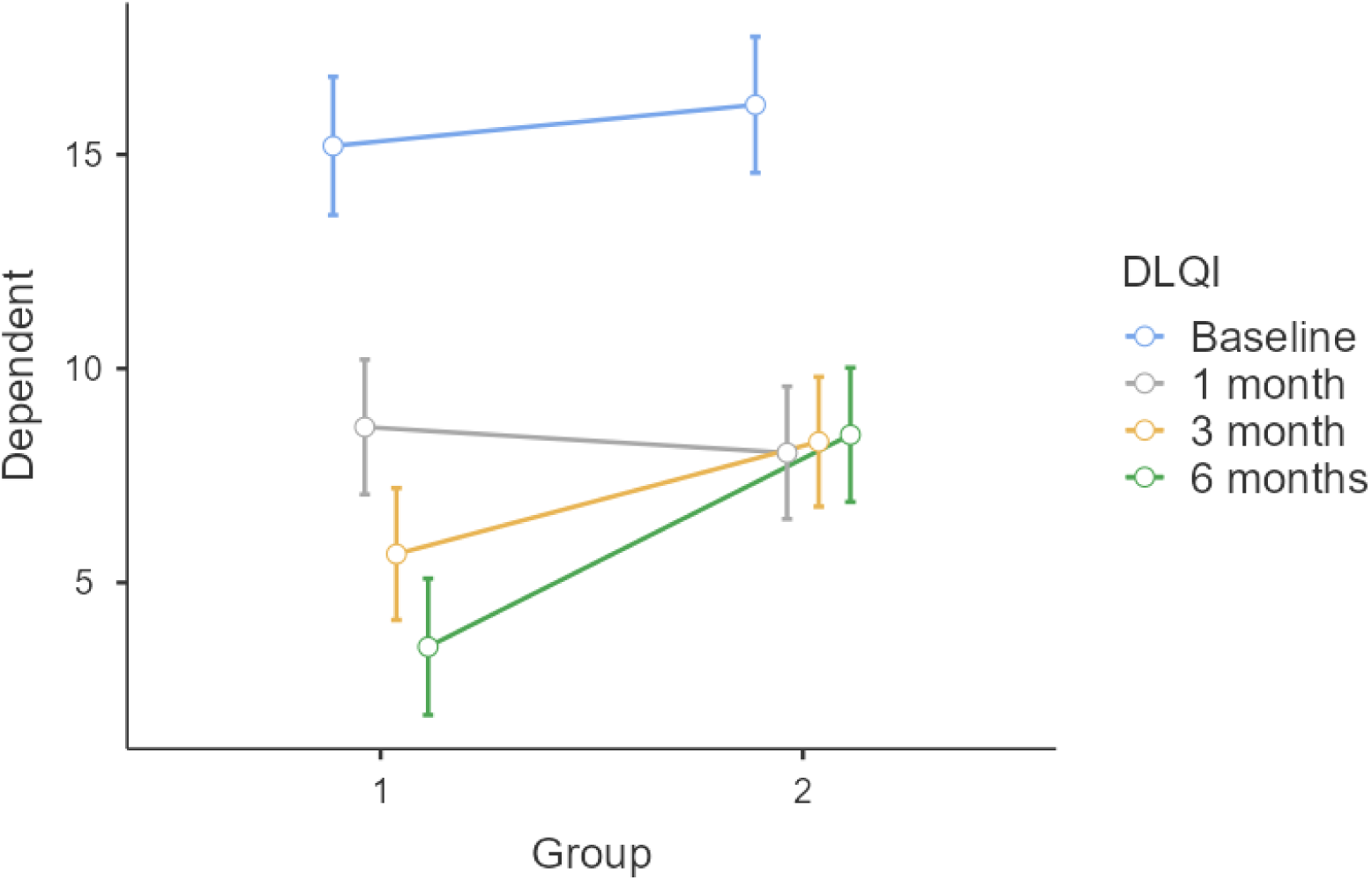
Comparison of Dermatology Life Quality Index between Group A and Group B during the follow-up period at 1 month, 3 months and 6 months.

Additionally, Spearman’s rank correlation analysis demonstrated a statistically significant positive correlation between the Neutrophil–Lymphocyte Ratio (NLR) and the ENLIST ENL Severity Score (EESS) at 6 months of follow-up (Spearman’s ρ = 0.269, degrees of freedom = 59, p = 0.018) as shown in Figure 7.

**Figure 7.**
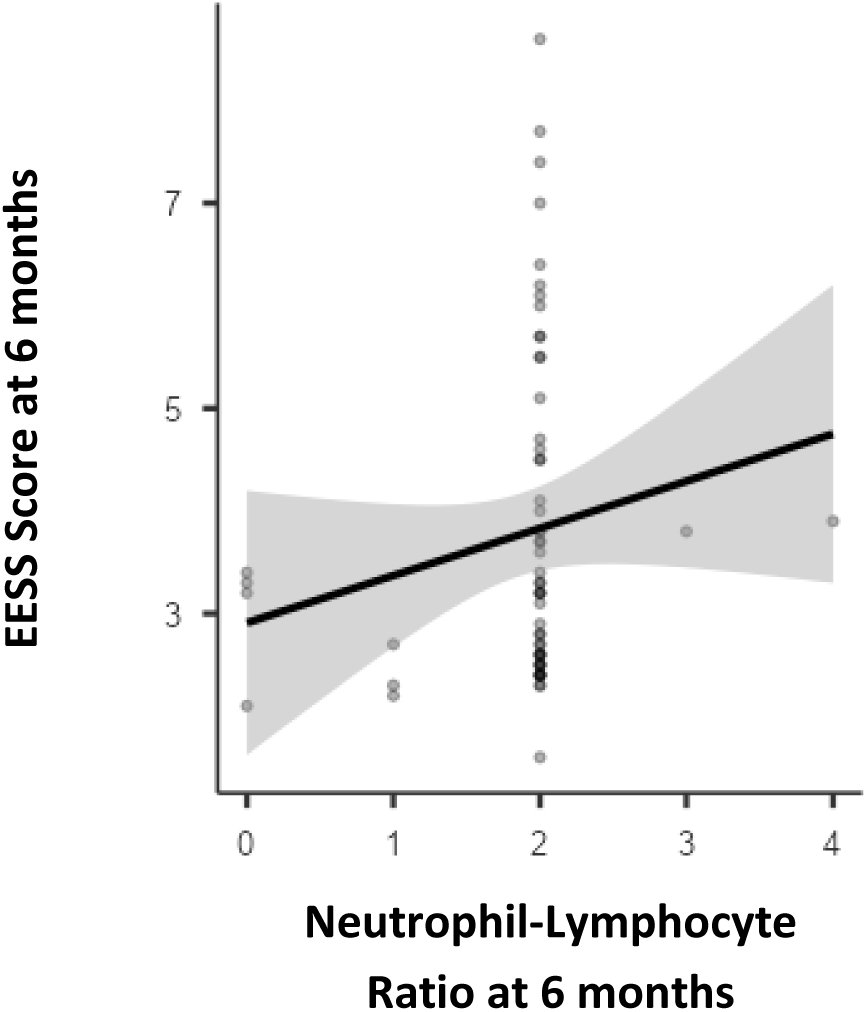
Spearman Correlation Analysis of NLR AND EESS Score at 6 months.

This finding indicates that higher NLR values were associated with higher disease severity scores at the end of follow-up. The direction of the association supports the alternative hypothesis (Hₐ) of a positive correlation between systemic inflammatory burden and ENL severity.

Although the strength of the correlation was modest, it remains clinically relevant, suggesting that persistent systemic inflammation contributes to ongoing disease activity. These results further reinforce the role of inflammation in influencing disease persistence and outcomes, and highlight NLR as a potential adjunctive biomarker for monitoring disease severity during follow-up.

Spearman’s rank correlation analysis demonstrated a moderate to strong positive correlation between Neutrophil-Lymphocyte Ratio (NLR) and Dermatology Life Quality Index (DLQI) at 6 months of follow-up (ρ = 0.604, p < 0.001) as shown in Figure 8.

**Figure 8.**
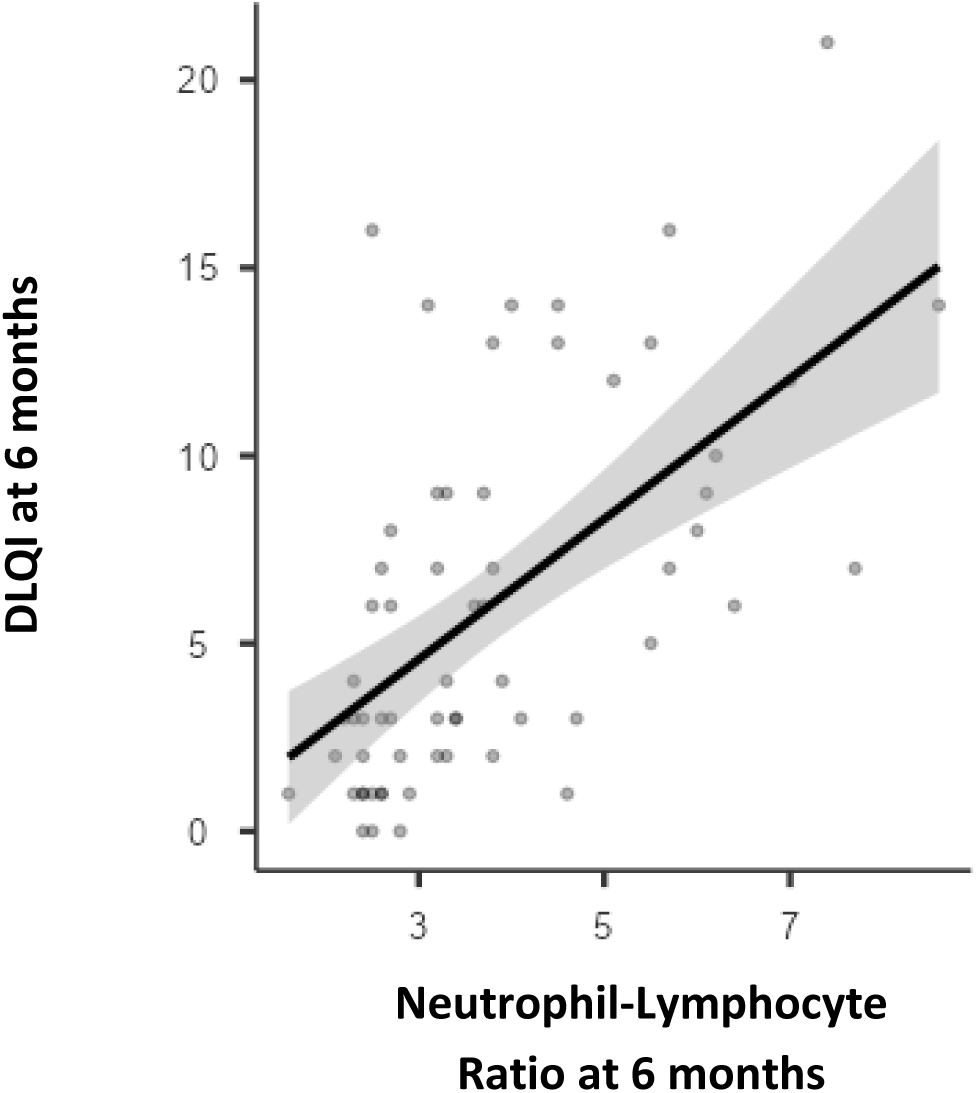
Spearman Correlation Analysis of NLR AND DLQI Score at 6 months.

This statistically significant correlation indicates that a higher systemic inflammatory burden, as reflected by elevated NLR values, was associated with worse dermatology-specific quality of life at the end of follow-up. Conversely, patients with lower NLR values tended to have lower DLQI scores, suggesting improved disease control and better patient-reported outcomes.

### Multivariable Logistic Regression Analysis for Relapse

Binary logistic regression was performed to identify independent predictors of relapse. Variables included treatment group, age, sex, baseline EESS score, baseline NLR, baseline DLQI score, duration of MDT completed at presentation, and socioeconomic status. Odds ratios (OR) represent adjusted odds after controlling for all variables included in the model. Statistical significance was defined as p<0.05.

Multivariable logistic regression analysis identified treatment group and baseline EESS score as significant independent predictors of relapse as shown in Table 3. Patients receiving prednisolone plus tofacitinib had significantly higher odds of relapse compared with those receiving prednisolone plus thalidomide (OR=9.77, 95% CI: 2.11–45.16, p=0.004). Thus, treatment with prednisolone plus tofacitinib was associated with nearly a ten-fold increase in the odds of relapse. Baseline EESS score was also significantly associated with relapse (OR=1.23, 95% CI: 1.01–1.48, p=0.037). Each one-unit increase in baseline EESS score increased the odds of relapse by approximately 22.6%.

**Table 3.** Multivariable binary logistic regression analysis evaluating factors associated with relapse among patients with Type 2 lepra reaction (ENL).

| Model Coefficients - Relapse |  |  |  |  |  |
| --- | --- | --- | --- | --- | --- |
|  |  |  | 95% Confidence Interval |  |  |
| Predictor | Estimate | Odds ratio | Lower | Upper | p |
| Intercept | -7.6027 | 4.99E-04 | 5.18E-07 | 0.481 | 0.03 |
| Group | 2.2789 | 9.766 | 2.1118 | 45.165 | **0.004** |
| Age (in years) | 0.0187 | 1.019 | 0.955 | 1.087 | 0.571 |
| Sex: |  |  |  |  |  |
| Male – Female | -0.2617 | 0.77 | 0.1496 | 3.961 | 0.754 |
| EESS (Baseline) | 0.2034 | 1.226 | 1.0127 | 1.483 | **0.037** |
| NLR (Baseline) | -0.1763 | 0.838 | 0.6862 | 1.024 | 0.084 |
| DLQI (Baseline) | 0.118 | 1.125 | 0.9453 | 1.339 | 0.185 |
| Months of MDT completed by patient at presentation (None, <1 year and > 1year): |  |  |  |  |  |
| 1 – 0 | 1.4208 | 4.14 | 0.7422 | 23.095 | 0.105 |
| 2 – 0 | -0.7687 | 0.464 | 0.0717 | 2.999 | 0.42 |
| Socio Economic Status: |  |  |  |  |  |
| Lower Middle – Lower | -0.9154 | 0.4 | 0.0296 | 5.421 | 0.491 |
| Upper Lower – Lower | -1.2279 | 0.293 | 0.0264 | 3.246 | 0.317 |
| Upper Middle – Lower | 3.4371 | 31.096 | 0.5566 | 1737.332 | 0.094 |
| Note. Estimates represent the log odds of "Relapse = 1" vs. "Relapse = 0" |  |  |  |  |  |

Age (OR=1.02, p=0.571), sex (OR=0.77, p=0.754), baseline NLR (OR=0.84, p=0.084), baseline DLQI (OR=1.13, p=0.185), duration of MDT completed at presentation (p>0.05), and socioeconomic status (p>0.05) were not significantly associated with relapse after adjustment for potential confounders.

### Adverse Effects Profile

No serious adverse events were observed in either group that necessitated hospitalization or permanent discontinuation of therapy during the study period. Overall, both treatment arms were well tolerated, with adverse events being predominantly mild to moderate in severity.

Constipation was significantly more frequent in Group A, reported in 30% of instances compared to 11.8% in Group B (p < 0.002). In contrast, gastrointestinal adverse effects, including diarrhoea, nausea, and abdominal pain, were significantly more common in Group B, occurring in 26% of instances, compared to 4.5% in Group A (p < 0.001).

Headache, drowsiness, and somnolence were observed more frequently in Group A (16.7%) than in Group B (2.5%), and this difference was statistically significant (p = 0.003).

Biochemical adverse effects, namely transaminitis and hyperlipidaemia, were significantly higher in Group B, reported in 28% of patients, compared to 3.6% in Group A (p < 0.001).

Minor upper respiratory symptoms, such as congestion of throat and nasal discharge, were reported sporadically in both groups without any statistically significant difference. Additionally, two patients in Group B developed herpes zoster during the 3-month follow-up visit. No long-term sequelae were observed in these patients.

## Discussion

Type 2 lepra reaction, also known as erythema nodosum leprosum (ENL), is a severe immune-mediated inflammatory complication that predominantly affects patients with multibacillary leprosy. Clinically, ENL presents with recurrent crops of painful erythematous nodules accompanied by systemic manifestations such as fever, neuritis, arthritis, lymphadenopathy and occasionally visceral involvement. Immunologically, ENL is a type III hypersensitivity reaction characterized by immune complex deposition, neutrophil activation and dysregulated cytokine signalling including Tumor necrosis factor-α (TNF-α), interleukin-6 (IL-6), interleukin-17 (IL-17) and interferon-γ. ^12^ These inflammatory pathways contribute to the chronic and relapsing nature of the disease.

Systemic corticosteroids remain the cornerstone of treatment for moderate to severe ENL because of their rapid anti-inflammatory effect. However, ENL often follows a chronic or recurrent course requiring prolonged corticosteroid therapy. Long-term steroid use is associated with substantial adverse effects including infections, metabolic disturbances, osteoporosis and adrenal suppression. Systemic corticosteroids have been highlighted as a “double-edged sword” in ENL management since prolonged dependence can lead to considerable morbidity. ^13^ Hence there has been always need for effective steroid-sparing agents in managing ENL. Among currently available therapies, Thalidomide remains the most effective drug for ENL owing to its potent anti-TNF-α activity and immunomodulatory effects. Thalidomide suppresses TNF-α production, modulates T-cell responses and reduces neutrophil migration, thereby interrupting the inflammatory cascade responsible for ENL.

In a randomized controlled trial from the Philippines, *Villahermosa et al.* evaluated 22 patients with ENL and found that thalidomide produced rapid clinical improvement in 86% of patients by Day 7, although most patients relapsed following discontinuation after 6 weeks, highlighting its efficacy for acute control but limited durability without maintenance.^14^ In a randomized trial by Kaur et al. involving 60 Indian patients with ENL, thalidomide (300 mg/day tapered over ∼6–8 weeks) produced faster clinical resolution and significantly lower relapse rates (∼13–15% vs >50%) compared to prednisolone (40 mg/day tapered over ∼10–12 weeks) during 6-month follow-up, demonstrating its superior efficacy in achieving sustained disease control. ^15^ Because of consistent efficacy reported in various studies, thalidomide is widely regarded as the gold-standard therapy for recurrent or chronic ENL despite its limitations such as teratogenicity, peripheral neuropathy and thromboembolic complications.

In a11-year retrospective cohort study from Hyderabad, India on 481 multibacillary leprosy patients *Pocaterra et al*., reported an ENL prevalence of 24%, with a strong association with lepromatous disease (49.4%) and high bacillary load. ENL predominantly affected middle-aged males (mean 34.7 years) and followed a recurrent course (mean 3.1 episodes over 18.5 months), with chronic ENL comprising 62.5% of cases. ^16^ These findings underscore the chronic, relapsing nature of ENL and its strong correlation with bacillary burden, emphasizing the need for prolonged and tailored management strategies.

Similarly, an Ethiopian cross-sectional study on clinic-epidemiological profile of 99 patients with ENL by Walker et.al, demonstrated a male predominance (61.8%) and high proportion of chronic ENL (70%) cases in the cohort. They highlighted that 74.5% cases of ENL presented in the early years within 1 year of initiation of multi-drug therapy. ^17^

The present study cohort was demographically and clinically similar to previous studies of Pocaterra et.al, and Walker et al., with a male predominant population (80.3%) of young to middle aged adults (mean age 35 years) and majority from rural and lower socio-economic background.

In a study by Nabarro et al., which was a retrospective audit conducted at the Hospital for Tropical Diseases, London, over a 17-year period (1996–2013), included 30 patients with ENL. The cohort demonstrated a high median bacillary index of 4.65, with 67% of patients developing ENL during MDT and 57% exhibiting a chronic disease course. ^18^ The median duration of ENL was 60 months (range 9–192), significantly longer in patients with BI > 4.5. Thalidomide was also administered in 87% of patients for a median duration of 16 months, and enabled steroid withdrawal within 2 months in 77% of cases, although 65% reported side effects. No deaths or thromboembolic events were observed. Jadhav et.al in a cross-sectional study in the year 1990 on 90 ENL patients reported, 100% patients to be in lepromatous pole (BL or LL). ^19^

Consistent with previous study of Nabarro et.al, 83.6% of our patients had BI>4+. In our study, 100% of patients were in Lepromatous pole which was similar to studies by Kaur et.al (95%) and Jadhav et. al (100%) but differing from Pocaterra et.al (49.4%). Additionally, recurrent ENL was the most common manifestation in our subjects (62.5%) as compared to other study by Walker et.al, and Pocaterra et.al respectively, who reported chronic ENL to be more common (70% vs 62.3%). ^16–19^ This observation is consistent with previous reports demonstrating a strong association between ENL and multibacillary disease.

In a 5-year record-based analysis of 102 Indian patients, Upputuri et al. demonstrated efficacy of Thalidomide in ENL patients. 66.7% were compliant with therapy and showed clinical improvement, with a relatively low recurrence rate of 16.2% and higher recovery rates observed in patients with lower bacillary index (≤4), acute ENL presentation, and those who had completed MDT. ^20^ Notably, patients completing MDT had a 2.5-fold higher likelihood of improvement. Thalidomide was generally well tolerated, with pedal oedema being the most common adverse effect (73.5%), and effective contraceptive counselling minimized teratogenic risk. Overall, the study highlighted that early initiation of thalidomide can induce faster remission, reduce recurrence, and can be safely implemented in outpatient settings with appropriate monitoring

In another report by, **Lockwood et al.** summarised rapid resolution of ENL lesions and significant reduction in relapse frequency among patients treated with thalidomide.^21^In the thalidomide arm of our study, patients demonstrated significant improvement in disease severity, inflammatory markers and quality-of-life indices.

The relapse rate in the thalidomide arm of our study was 36.7 % over the 6-month follow-up period but 93.3% were weaned off steroids after 6 months of treatment. The mean relapse burden was significantly lower when compared with the tofacitinib group. These findings are broadly consistent with previous studies by Kaur et.al (15.2% relapse) and Upputri et.al (16% relapse).

NLR has increasingly been recognized as a marker of systemic inflammation. The **progressive reduction in neutrophil-lymphocyte ratio (NLR)** in patients receiving thalidomide or tofacitinib was an important finding in our study. A study by Tonojo et al. conducted on 182 patients with multibacillary leprosy, including 22 cases of ENL, demonstrated that haematological ratios such as NLR and PLR have significant diagnostic value in ENL. The study reported that NLR, at a cut-off value of 4.99, showed high sensitivity (86.4%) and specificity (82.5%), indicating strong diagnostic accuracy for identifying ENL. ^22^ Schimtz et.al, from Brazil in a report proposed possible role of NLR in leprosy patients as marker of inflammation based on the underlying pathogenesis. ^23^ In our study, NLR demonstrated a significant positive correlation with ENLIST ENL Severity Score and Dermatology Life Quality Index, suggesting that systemic inflammatory burden is closely related to disease severity and patient-reported outcomes.

In recent years, targeted immunomodulatory therapies have been explored for inflammatory dermatological diseases. **Tofacitinib**, an oral Janus kinase (JAK-1 and JAK-3) inhibitor, modulates intracellular signalling pathways involved in cytokine-mediated inflammation. By inhibiting the JAK-STAT pathway, tofacitinib suppresses several cytokines implicated in ENL pathogenesis including IL-6, IL-17, TNF-α and interferon-γ.^23^

Although evidence remains limited, a few case reports have suggested potential benefit of tofacitinib in lepra reactions. **Patel et al.,** reported a case of Lucio phenomenon associated with dual infection by Mycobacterium leprae and Mycobacterium lepromatosis in which adjuvant tofacitinib therapy resulted in dramatic clinical improvement with resolution of ulcerative lesions.^24^ In another Indian case report **by Benaka et.al**, reported clinical improvement in recurrent Type 2 lepra reaction refractory to Thalidomide and systemic corticosteroids in a 31 year old male patient prescribed with Tofacitinib 11mg extended release tablet. ^25^ Similarly, **Nakamura et al.** used Tofacitinib in patients of ulcerative colitis developing erythema nodosum and achieved significant response in resolution of the inflammatory lesions. ^26^Through this report, they proposed the intracellular signalling pathway and highlighted the role of Janus Kinase inhibitors (JAK) in modulating cytokine derived inflammation. These observations suggest that JAK inhibitors may modulate inflammatory pathways involved in lepra reactions.

In our study, although patients receiving tofacitinib demonstrated initial improvement in disease severity, relapse occurred in 71 % of patients in this group, which was significantly higher than the thalidomide group. Furthermore, improvement in quality-of-life scores plateaued during follow-up, and systemic inflammatory markers remained relatively elevated compared with the thalidomide group.

The present study therefore provides important comparative evidence evaluating **tofacitinib versus thalidomide as adjuvant therapy to corticosteroids in ENL**.

In addition, current study compared both drugs in terms of disease severity, relapse, NLR and DLQI improvement. Although both drugs were effective in treatment of ENL, Thalidomide had better response when compared to Tofacitinib in terms of relapse rate (36.7% vs 71%), patients off steroid (93.3% vs 58.7%) and decline in NLR (2.75 vs 4.50). Although EESS decreased significantly at end of 6 months follow-up in both groups but the response plateaued at 3 months in the latter. Taken together, these findings suggest that while tofacitinib may exert immunomodulatory effects through inhibition of cytokine signalling, its efficacy as a steroid-sparing therapy appears relatively less promising to thalidomide within the six-month follow-up period of the present study. Thus, Tofacitinib could be a plausible alternative adjuvant with cautious monitoring with fewer side-effects and scenarios where Thalidomide is contraindicated.

Hence the present study adds to the existing literature by providing **one of the first comparative clinical analyses evaluating tofacitinib against thalidomide in the management of ENL**.

Another notable finding of our study was the correlation between NLR and ENL severity, suggesting that NLR may serve as a simple and accessible biomarker for monitoring inflammatory activity and treatment response during follow-up.

Despite these important findings, several limitations should be acknowledged. The study was conducted at a single tertiary-care centre with a relatively small sample size and limited follow-up duration. In addition, the observational design with a historical comparison group may introduce potential bias. Larger multicentric prospective studies and randomized controlled trials will be required to validate these findings and further clarify the role of targeted immunomodulators in ENL.

In conclusion, the present study demonstrates that **thalidomide holds better efficacy to tofacitinib as an adjuvant therapy to corticosteroids in patients with ENL**, particularly with regard to relapse prevention, steroid withdrawal and sustained improvement in disease severity and quality of life. However, Tofacitinib could be an alternative option in management recurrent and chronic ENL whenever the former is contraindicated, not tolerated or unaffordable. There were various morphologies of ENL that we have encountered in this study as shown in Figure 9a-9g

The study also highlights the potential utility of neutrophil-lymphocyte ratio as a biomarker for disease activity. Future research should focus on larger controlled trials evaluating targeted immunomodulators such as JAK inhibitors and biologic agents in order to expand therapeutic options for this challenging complication of Hansen disease.

**Figure 9a.**
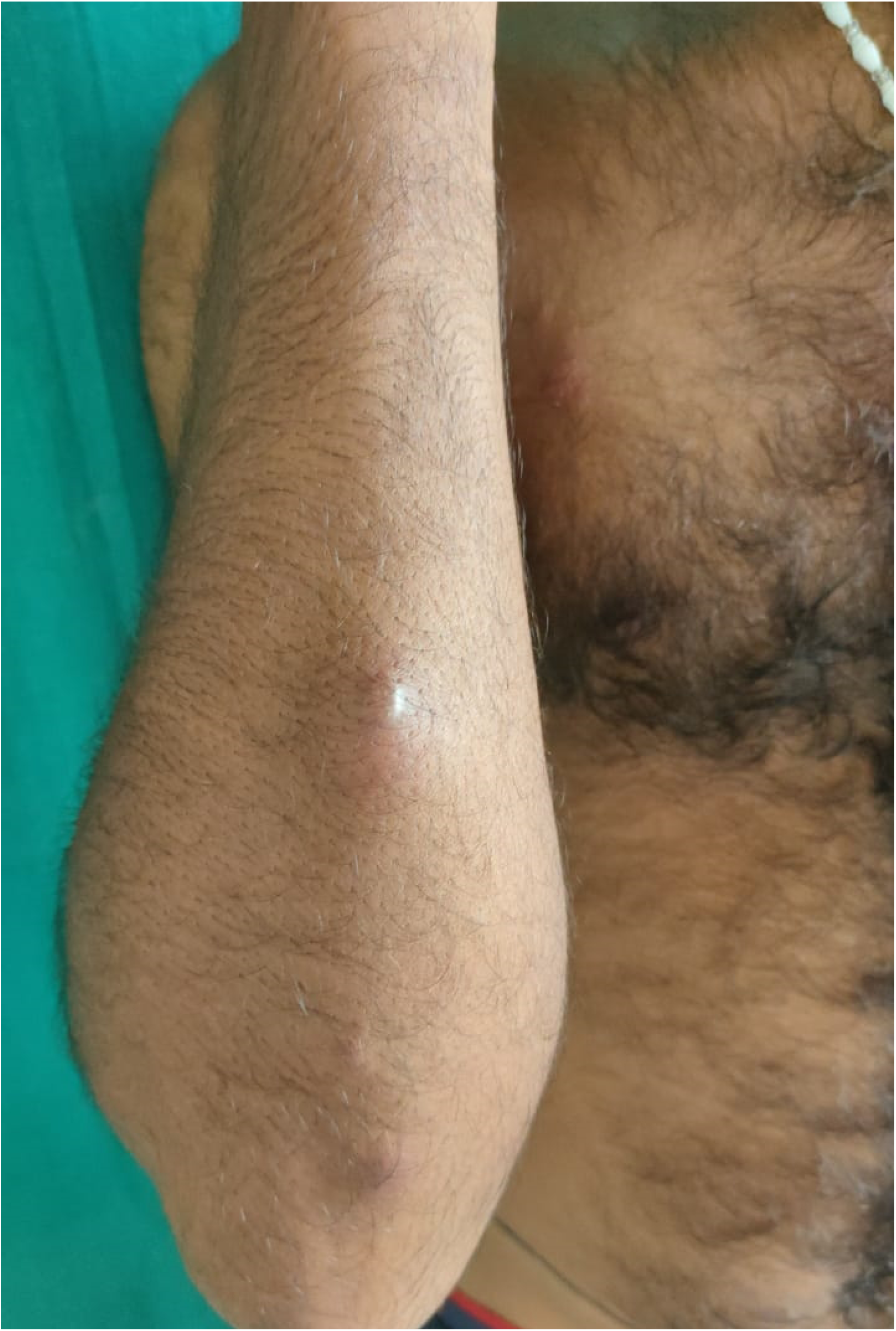
One of the study subject having Bullous ENL lesion on the arm.

**Figure 9b.**
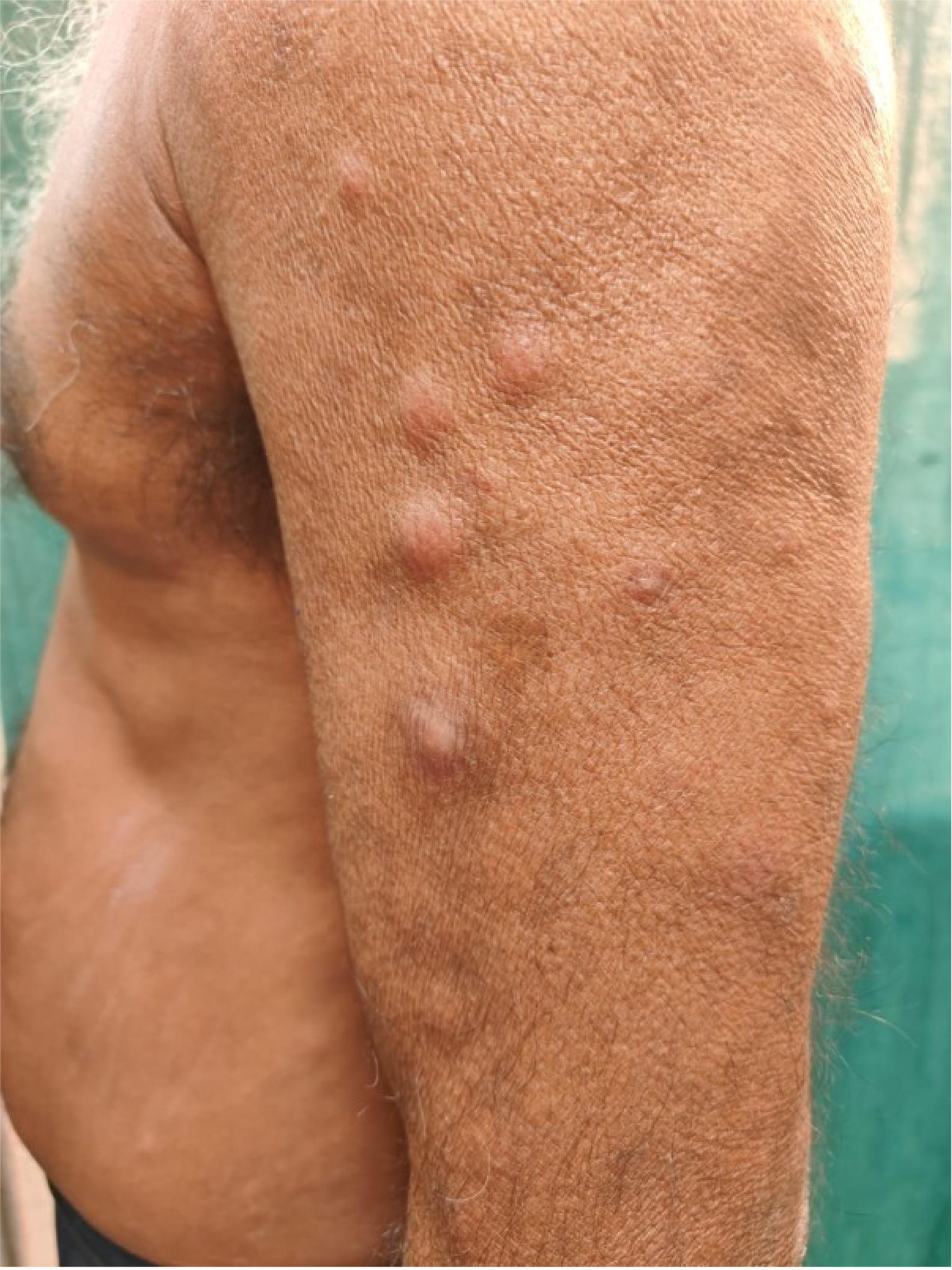
One of the study subject having classical Nodular ENL lesion on the arm.

**Figure 9c.**
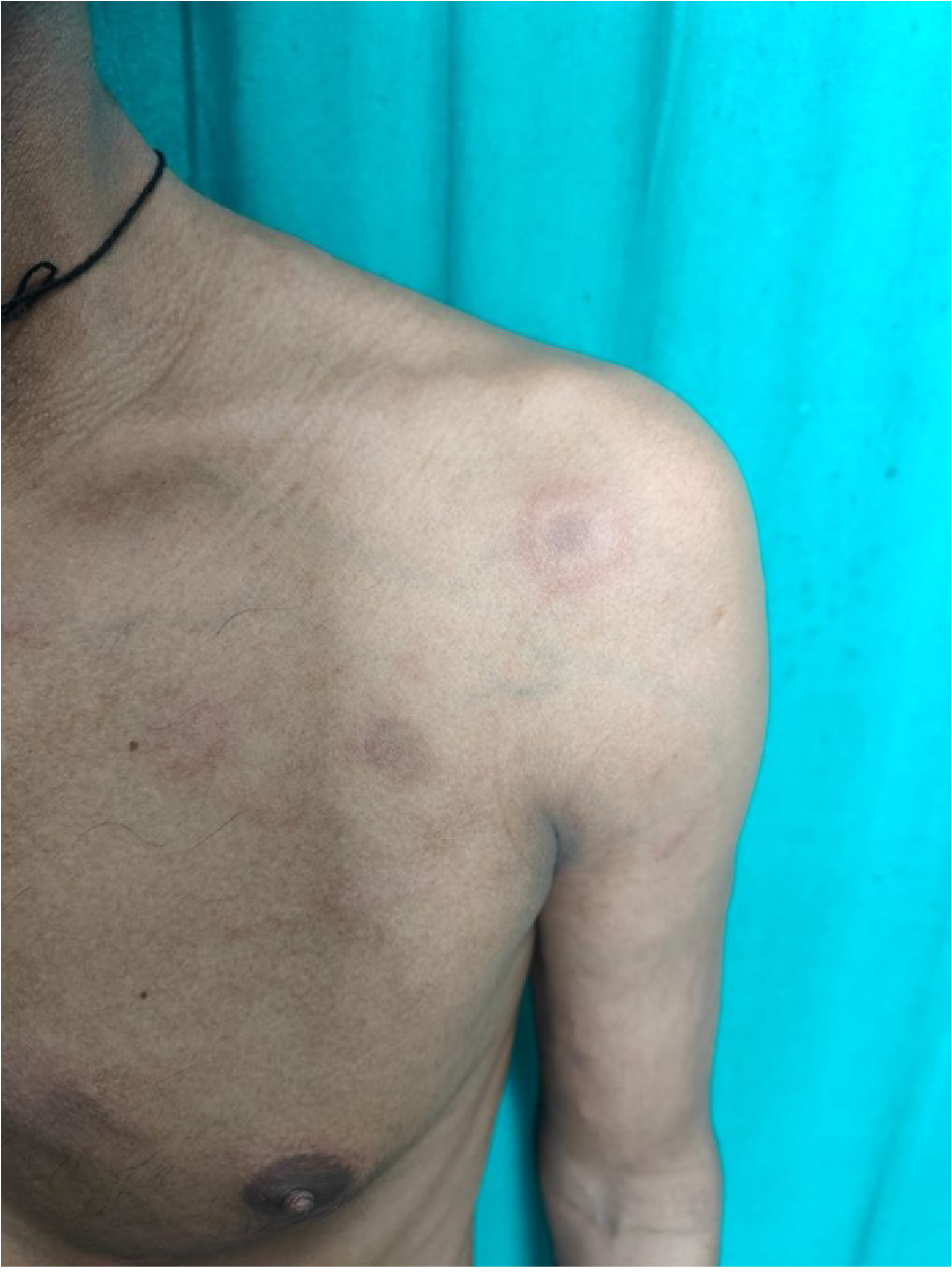
One of the study subject having Erythema Multiforme like ENL lesion on the arm.

**Figure 9d.**
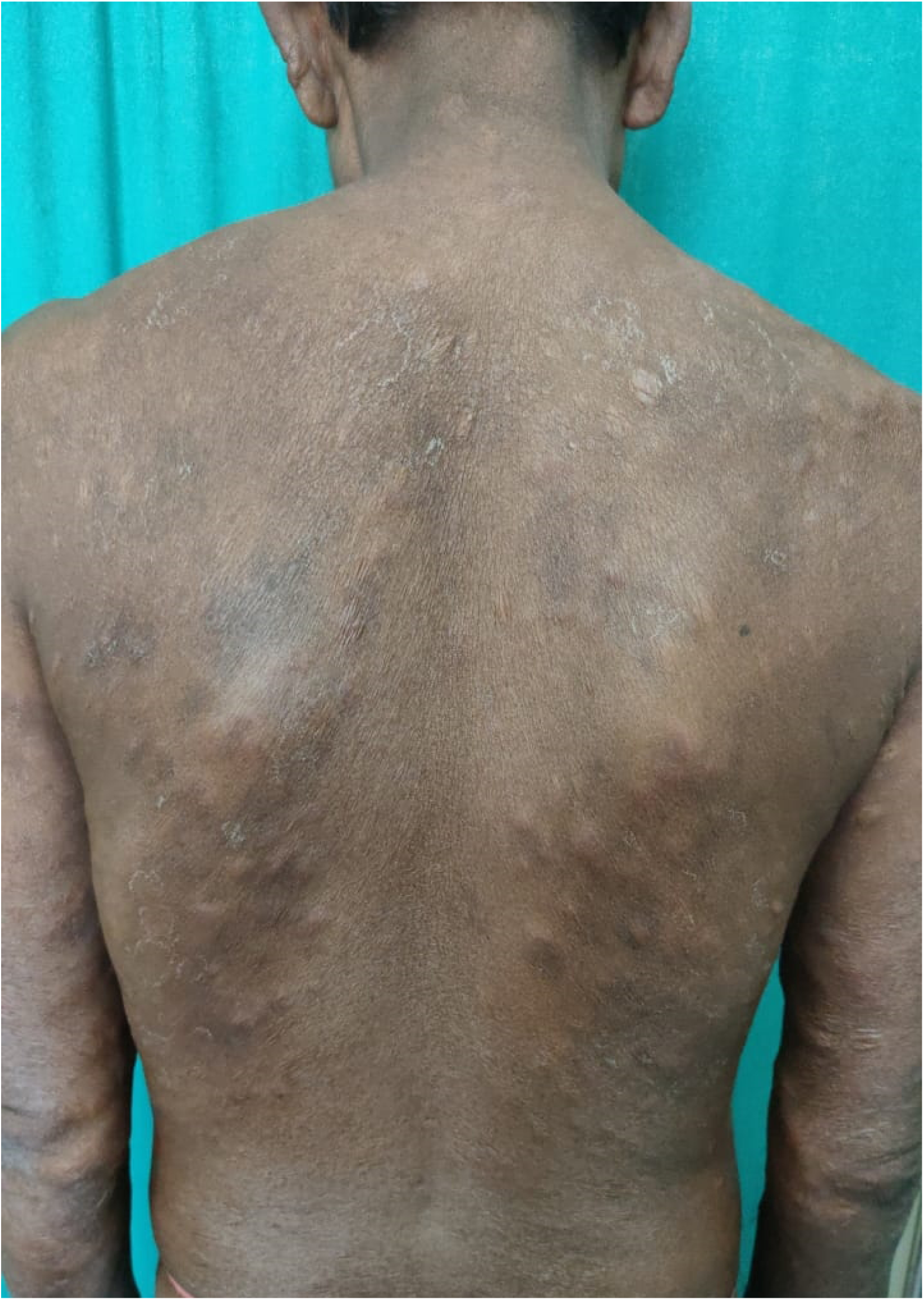
Extensive distribution of ENL over the back as nodular lesion, scars and many healing with post inflammatory hyperpigmentation.

**Figure 9e.**
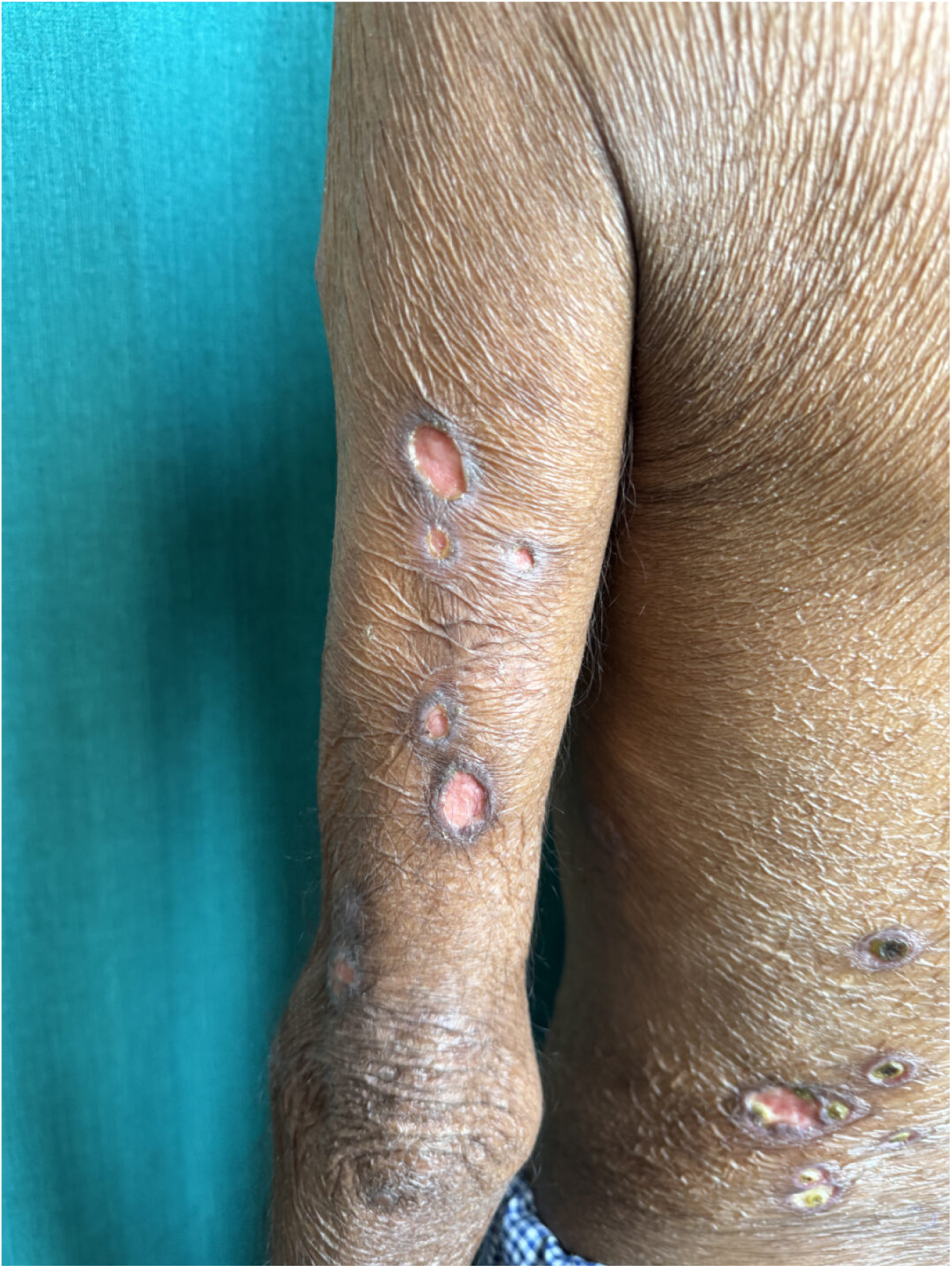
One of our study subject having Necrotic ENL on the forearm and back.

**Figure 9f.**
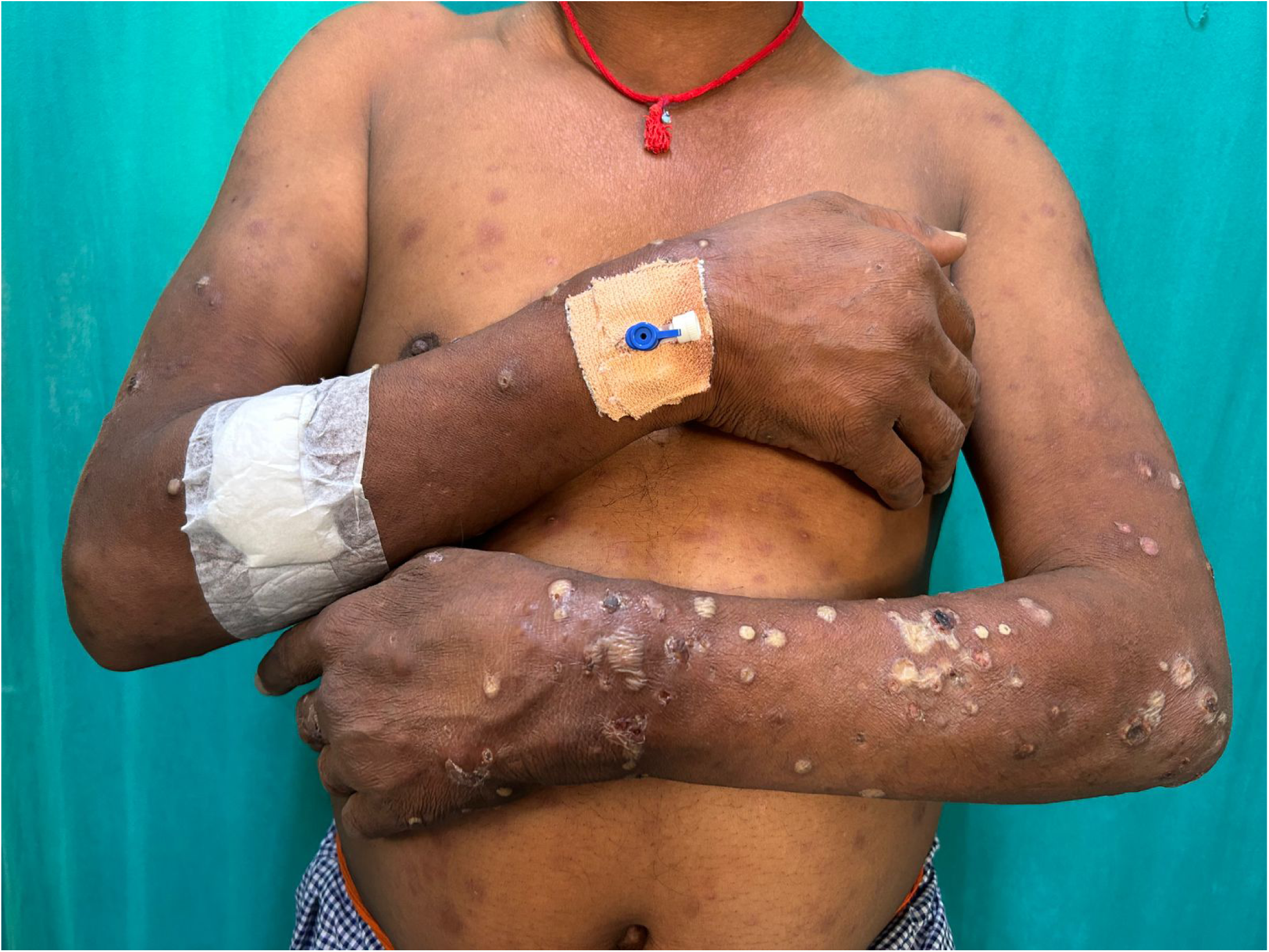
One of our study subject having Pustular ENL on the forearm.

**Figure 9g.**
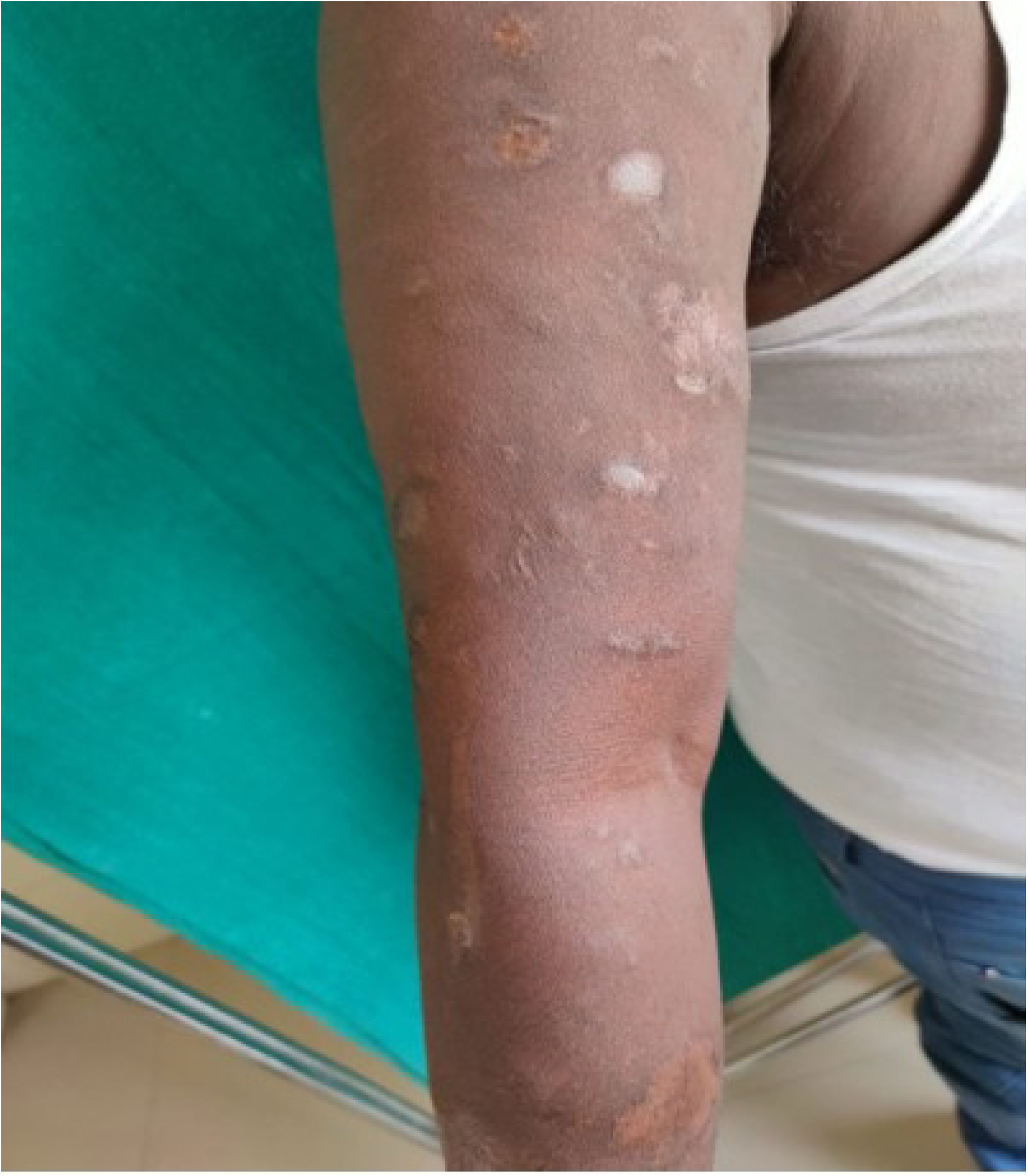
One of our study subject having resolved necrotic ENL on the forearm – resolving with atrophic scar.

## Data Availability

Data may be available to the readers on request on the discretion of the authors

